# Migrant healthcare workers during COVID-19: bringing an intersectional health system-related approach into pandemic protection. A German case study

**DOI:** 10.1101/2023.01.28.23285135

**Authors:** Ellen Kuhlmann, Marius-Ionut Ungureanu, Georg MN Behrens, Anne Cossmann, Leonie Mac Fehr, Sandra Klawitter, Marie Mikuteit, Frank Müller, Nancy Thilo, Monica Georgina Brînzac, Alexandra Dopfer-Jablonka

## Abstract

**Introduction:** Migrant healthcare workers played an important role during the COVID-19 pandemic, but data are lacking especially for high-resourced European healthcare systems. This study aims to research migrant healthcare workers through an intersectional health system-related approach, using Germany as a case study.

**Methods:** An intersectional research framework was created and a rapid scoping study performed. Secondary analysis of selected items taken from two COVID-19 surveys was undertaken to compare perceptions of national and foreign-born healthcare workers, using descriptive statistics.

**Results:** Available research is focused on worst-case pandemic scenarios of Brazil and the United Kingdom, highlighting racialised discrimination and higher risks of migrant healthcare workers. The German data did not reveal significant differences between national-born and foreign-born healthcare workers for items related to health status including SARS-CoV-2 infection and vaccination, and perception of infection risk, protective workplace measures, and government measures, but items related to social participation and work conditions with higher infection risk indicate a higher burden of migrant healthcare workers.

**Conclusions:** COVID-19 pandemic policy must include migrant healthcare workers, but simply adding the migration status is not enough. We introduce an intersectional health systems-related approach to understand how pandemic policies create social inequalities and how the protection of migrant healthcare workers may be improved.

## Introduction

The migrant healthcare workforce (HCWF) is an important pillar of the health labour market (1). During the COVID-19 pandemic, migrant healthcare workers (HCWs) played a crucial role in maintaining healthcare delivery and resilience of the health system (2, 3). This was especially relevant in high-income European countries that suffer from chronic health labour market shortages and increasingly depend on foreign health human resources (4, 5). In 2020, Germany, Spain and the United Kingdom were the main destination countries in the WHO European Region for foreign-trained physicians and nurses, in absolute numbers (1). In Germany, for instance, 13.8% of physicians and 9.2% of nurses were foreign-trained (6).

Despite their health labour market role, the migrant HCWF remained largely invisible in the pandemic policy discourse. Epidemiological data (7), as well as health labour market demand and individual needs of migrant HCWs, were poorly reflected in national pandemic protection plans and global health programs (8). Even three years after the COVID-19 outbreak, systematic monitoring is still lacking. However, research revealed an overall exacerbation of social inequalities in societies during the COVID-19 pandemic and higher rates of infection, hospitalisation, or death among migrants and ethnic minorities (9, 10). This was reported from high-income countries (11), including those with well-established universal health coverage and social inclusive healthcare systems in the European Union (EU), like Denmark (12). Across the globe, there is also mounting evidence that the pandemic increased gender inequalities and hit women the most (13, 14), including women HCWs (15, 16). Notably, women account for the vast majority of HCWs in all healthcare systems (1).

Concerning the health workforce, there is a dearth of knowledge on migrant HCWs, but some information became available recently from research into social inequalities that considers race/ethnicity. A Brazilian survey confirmed an exacerbation of essential inequalities among HCWs during the pandemic and highlighted intersections between race, gender and profession that may affect HCWs in different ways (17, 18). In the United Kingdom (UK), the United Kingdom Research study into Ethnicity and COVID-19 outcomes among Healthcare workers (UK Kingdom-REACH) (19, 20, 21) and efforts undertaken by the British Medical Association (22) revealed strong racial inequalities and higher risks of migrant HCWs. COVID-19 hit HCWs with Black, Asian and Minority Ethnic (BAME) backgrounds most strongly, thus putting a spotlight on structural racism in the British National Health Service (NHS) (23).

However, data were mainly collected in countries that were strongly affected by the COVID-19 pandemic, struggling with underfunded healthcare systems and poor pandemic management, often caused by populist governments that denied the threats of COVID-19 (24, 25, 26). Comparative information gathered in a broader range of health systems with more effective pandemic policy is missing. Data are especially poor for the EU region. This also raises questions about the situation in countries with a well-resourced healthcare system and hospital sector, like Germany (27, 28, 29), that managed the COVID-19 pandemic comparatively well (30). It is also not clear, how racialised discrimination reported from the UK and Brazil played out in the EU health labour market with its free movement and high cross-country mobility. Most importantly, the migrant status itself is no uniform category and a common definition is lacking. Comparison of data, where they exist, is therefore challenging. A recent WHO report identified the items ‘foreign-born’ and ‘foreign-trained’ as the most popular categories (1), whereas the OECD provides information on ‘foreign-trained’ physicians and nurses in relation to ‘domestically-trained’ (6).

Against this backdrop, we seek to close a gap and explore the situation of migrant HCWs through an intersectional lens in a health system with more effective pandemic measures. The German healthcare system and the organisational setting of Hannover Medical School, characterised by a comparably low infection risk of HCWs (31) serve our analysis as an optimal-case scenario. Our study aims to contribute new knowledge to HCWF policy and to reveal how health system conditions may exacerbate existing inequalities during a global public health crisis.

## Methods

### Developing an intersectional research design

Theoretically, the research is informed by a health system approach (30), an intersectional perspective on social inequalities exacerbated by the COVID-19 pandemic, and a focus on actor-centred data (31). The connectedness of epidemiological and social conditions of the COVID-19 pandemic has been described in different ways. For instance, in a Lancet editorial, Horton (32) introduced the term ‘syndemic’ in the early stage of the crisis, but there is still little theoretical foundation and few authors utilised the concept (33, 34).

Advanced theoretical concepts are available from gender, race, and social inequality studies. Feminist researchers, in particular, were able to operationalise and empirically explore complex social dynamics during the COVID-19 pandemic through an intersectional lens (13, 15, 35). Characteristically, this research is sensitive to structural inequalities and the effects of pandemic policy and politics; see also the Solidarity in times of Pandemics Research Consortium (36). A few other studies provide useful hints on organisational and individual conditions that may have caused inequalities in the HCWF during the pandemic (37, 38, 39), but data are limited because they are either not disaggregated for migrant HCWs, or submerged into a category of ‘essential workers’.

Figure 1 introduces an intersectional health-system related framework that is guiding our research design and empirical analysis.

**Figure 1.**
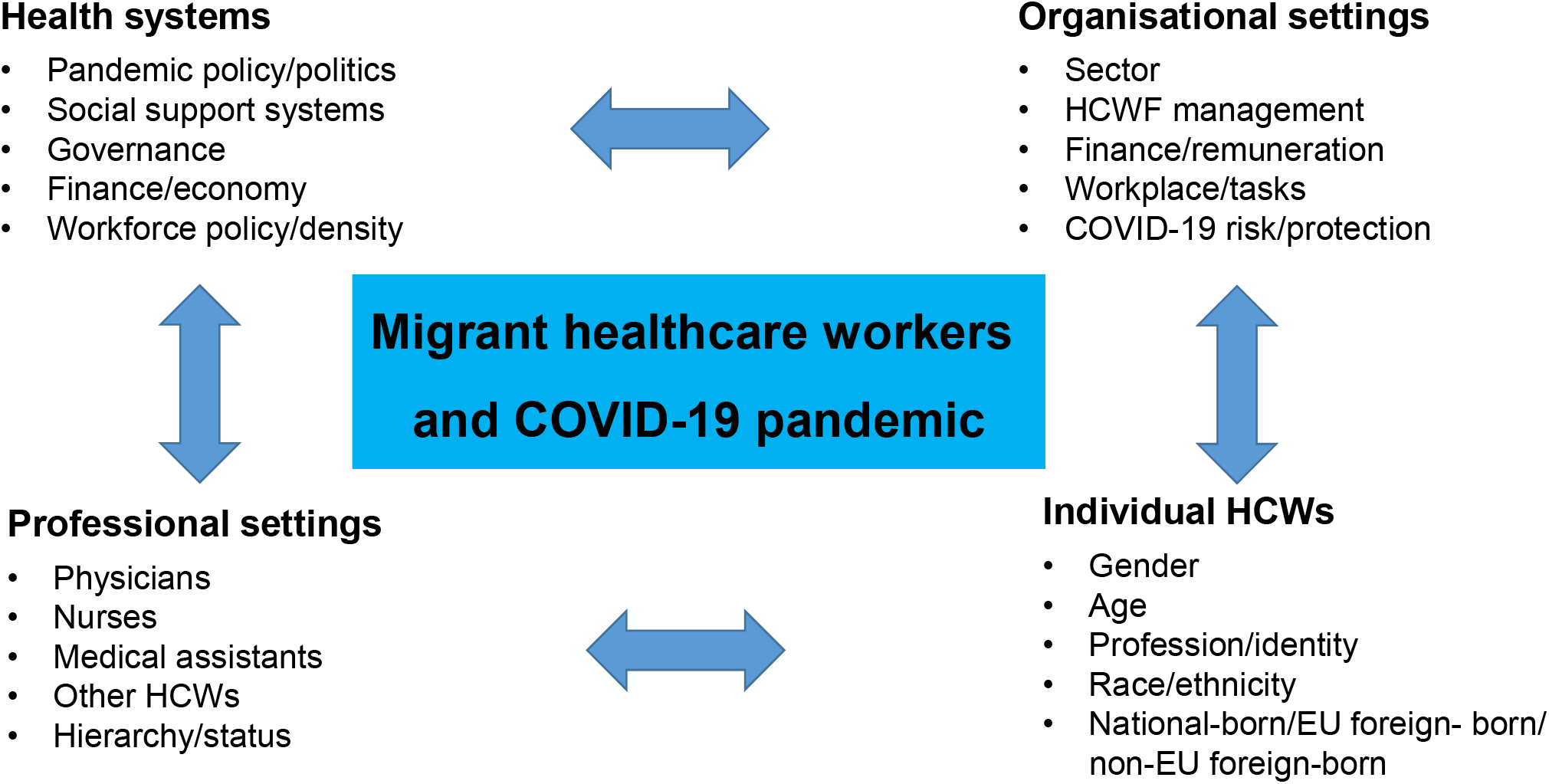
A Generic intersectional research framework nested in health systems and governance Source: authors’ own figure

To begin with, our German case study reflects a health system (27) with significantly lower excess mortality than in comparable regions (30) that was relatively well prepared in terms of infrastructures and resources, spent a greater proportion of its GDP on health (11.7%) than any other EU country, and was among the OECD countries with the highest health workforce staffing levels (8, 29, 30, 40), although workforce shortage was worsening. Further explanations for Germany’s more successful pandemic management include, among others, a high amount of testing, a lower median age of those infected and the highest number of intensive care beds per 100,000 people in Europe (30). Greer and colleagues (33) furthermore highlight the importance of the governance structure and an effective allocation of authority.

On the organisational level, we have chosen the hospital sector and a university hospital/ academic health centre located in the State (*Land*) of Lower Saxony in the Western part of Germany. The German hospital sector is generally well-equipped and additional measures were taken to ramp up infection control and protection of HCWs and, to a lesser degree, the psycho-social support of HCWs (41, 42, 43). As described elsewhere, ‘in 2020, when the CoCo Study was initiated, routine capacities of hospital beds accounted for 1,520 beds and 7,500 employees, including 3,100 HCWs (head counts)’ (31, 44).

Concerning the professional settings, our case study focuses on HCWs in the hospital sector, where the qualification of HCWs is overall high compared to the primary care and long-term care sectors; nurses are the largest and physicians the second largest group (27). There is also evidence that the positive effects of strong professionalism in higher-status groups may dominate identity concepts of HCWs, thus mitigating potentially negative experiences of migration status (45).

Against this backdrop, our selected research design mirrors an optimal-case scenario in relation to the system and the organisational and professional setting, while actor-centred perceptions (micro-level) of HCWs will be empirically analysed in a comparative perspective. We compare national-born and foreign-born HCWs, and if possible, further differentiate EU foreign-born and non-EU foreign-born, assuming that EU health labour market regulation and policy support for cross-national mobility during the pandemic (28) may create different conditions for these groups. These categories reflect the common practice of OECD (2) and WHO (1), as well as feasibility; data related to other categories are largely missing for Germany and ‘race’, in particular, is absent from public statistics and debate in Germany and rarely considered in European countries, except the UK.

### Data sources and secondary analysis

In our study, we applied an explorative approach and sought to combine different methods and data sources to respond to the challenges of the ‘HCW migrant’ category. Guided by the generic framework (Figure 1), a rapid scoping study of the international literature was carried out in November 2022 to gather empirical information on intersectional inequalities in the HCWF and place the German case in context. Subsequently, secondary analysis (46, 47) of data taken from two surveys carried out at Hannover Medical School was performed. This methodological approach seems to be most helpful to close essential knowledge gaps promptly, since primary data are lacking and no systematic monitoring has been established. It also contributes to the effective and ecological use of research resources (46, 47).

#### Scoping study

A rapid scoping review methodology was chosen, drawing on a qualitative approach (48). This approach recognised that data were limited and novel challenges were emerging during a major global health crisis, in addition to those of the migration category. The aim was to highlight gaps in the literature and identify conditions that may exacerbate social inequalities in the HCWF to order to inform further research and policy recommendations, rather than systematically analyse evidence. More specifically, PubMed searches were conducted (November 2022) combing the key related terms ‘healthcare workers’ and ‘COVID-19 pandemic’ according to the guiding framework to two major strands of the literature, namely ‘migrant’ and ‘social inequalities’. In addition, an extensive hand search was performed, including documents, websites, professional associations, and key research projects; conference contributions and expert information were also considered (Supplement Figure S1).

The main inclusion criterion was information on migrant HCWs with a professional background during the pandemic; exclusion criteria were carers, informal care and the long-term care sector, where the COVID-19 situation was markedly different (49, 50), as well as general information on migrants, studies on essential workers without specified data on HCWs, and research into migrant HCWs not related to COVID-19. Records with marginal information on our topic, as well as study protocols and mere opinion papers and editorials without substantive empirical data were also excluded. Finally, 19 articles were selected for the thematic analysis (17, 45, 51, 52, 53, 54, 55, 56, 57, 58, 59, 60, 61, 62, 63, 64, 65, 66, 67).

Three major coding categories were defined to extract information on migrant HCWs during the COVID-19 pandemic. Category 1 primarily sought to reveal *whether* information is available, while the other two categories explored the substance of inequalities comparatively.

1. Availability of data and inclusion of migrant HCWs in pandemic policy.
2. Comparison of social inequalities and the substance of disadvantages and discrimination.
3. Identification of intersectional inequalities and relevant categories.

The analysis followed a two-step approach. The first step was related to category 1 and summarised relevant information regardless of the source and article category, including expert information, websites, and documents (e.g. from professional associations) next to the selected articles. In step 2, thematic analysis of selected research and review articles (Figure 1A) was performed for coding categories 2 and 3.

- *Secondary analysis, DEFEAT Corona study and COVID-19 Contact (CoCo) study* The DEFEnse Against COVID-19 STudy (DEFEAT) (https://www.defeat-corona.de) is a digital Long Covid project related to the long-term effects of the corona pandemic with a focus on Lower Saxony, Germany (68). It is hosted at Hannover Medical School in collaboration with University Medicine Göttingen and Ostfalia University of Applied Sciences – Hochschule Braunschweig/Wolfenbüttel. In spring 2022, more than 5,000 persons participated in the survey (questionnaire available in German) (68). This data set served our analysis. It allowed disaggregated data for national-born and foreign-born HCWs, but no information was available on the profession and whether the country of origin belongs to the EU.

The selected items comprised health-related information (e.g. vaccination, SARS-CoV2 infection), a Quality of Life score (QoL EQ-5D; higher value=better QoL) (69), and a score for social participation (IMET; higher value=higher impairment of social participation) (70). In addition, we unpacked the IMET score and selected two single items: ‘social activities’ and ‘stress/burdens’. Descriptive statistical analysis was performed to compare the two HCW groups, using Chi-square or Fisher Exact test for categorical variables and Wilcox test for continuous variables.

The COVID-19 Contact (CoCo) (https://www.cocostudie.de/) study is a multi-method study gathered at Hannover Medical School that comprised SARS-CoV-2 serology testing of HCWs with patient contact in low-prevalence settings and a questionnaire-based online survey (44, 71). Inclusion criteria were all HCWs working at Hannover Medical School in patient care or in units with possible COVID-19 contact (German Clinical Trial Registry, DRKS00021152; study protocol: https://doi.org/10.1101/2020.12.02.20242479) (44). For our study, questionnaire data collected during the second wave of the COVID-19 pandemic in Germany, from November 2020 to February 2021, were selected. The initial questionnaire did not provide information on the migration status of HCWs. Yet the longitudinal design of the CoCo study included an anonymised tracking system of participants that allows connecting of different surveys over time. Thus, we were able to collect the information ex-post and create a sample for our comparison via hand search, differentiated for national-born, EU foreign-born, and non-EU foreign-born.

For our comparative analysis of migrant HCWs, overall 16 items (CoCo 2.0, questionnaire 2; in German) (44, 71) were selected:

- a description of the sample based on gender, age, workplace, profession, and children/childcare responsibilities (5 items);
- perceptions of individual, organisational and governmental issues related to COVID-19 (11 items).

Descriptive statistical analysis was undertaken to compare the three groups of HCWs. In a second step, the groups of EU foreign-born and non-EU foreign-born were merged and compared to national-born HCWs. Kruskal-Wallis test was performed for items related to the perceptions of HCWs; Pearson’s Chi-squared test with Yates’ continuity correction for gender and children/caring related items; Fisher’s Exact test for health status and workplace-related items, specified by Fisher’s Exact test with simulated p-value (based on 2000 replicates) to consider the EU and non-EU foreign-born groups for the workplace item.

## Results

### Intersectional inequalities of migrant HCWs in health system context: rapid scoping review

#### What data are available on migrant HCWs?

The scoping study put a spotlight on the lack of attention to migrant HCWs during the COVID-19 pandemic. Scarcity of knowledge was observed globally and nationally in Germany for both the health labour market and actor-centred data, despite significant investigations into health system data and monitoring (29). Recently, the HCWF is gaining greater recognition in global health policy and information is improving (1), yet data are rarely connected to migration and/or social inequalities (8, 82). This observation described in the literature is also true for Germany (38, 73, 74) and most other countries, as well as for major European (29, 75) and international databases. The OECD (2) performed a helpful health labour market analysis in relation to the contribution of the migrant HCWF during the pandemic, but did not consider the ‘human face’, the perceptions of individual HCWs (76). The WHO also moved the HCWF higher up on the agenda (1) and is scaling-up efforts in 2023.

Information provided by professional associations was also scarce. The International Council of Nurses (77) documented mobility flows during the pandemic, but did not consider cross-country comparison of migrant HCWs. In Europe, the Standing Committee of European Doctors (78, 79) sought to gather data, yet members could not provide the information, except the British Medical Association (BMA) (22).

Knowledge of migrant HCWs during the pandemic is strongly shaped by two country clusters, Brazil (17, 18, 62, 63, 80) and the UK, the latter one mainly based on data from the UK-REACH study (20, 21) and efforts of the BMA (22, 23, 81, 82, 83, 84). Some additional information was provided by qualitative research with frontline Pakistani emigrant physicians in the National Health Service (NHS) as a specific group of BAME HCWs but did not include comparison (85). Further research was conducted in Sweden (66), in the USA (64) and Quebec, Canada (65). The Canadian and Swedish studies and a few reports of international associations considered migrant HCWs more explicitly (67, 77). In Germany, we found overall three studies with some information on migrant HCWs (45, 51, 52), but one was only marginally related to HCWs and another one not specifically concerned with the situation during the COVID-19 pandemic.

#### What information is available on inequalities and the substance of disadvantages?

Comparative data revealed strong social inequalities and discrimination against migrant HCWs and racialised groups in the HCWF. This was especially documented for BAME doctors in the UK (53, 54, 56, 57, 58, 59, 60) and ethnic minorities in Brazil (17, 62, 63). Black HCWs faced the strongest discrimination in most countries included in the analysis, as shown for the UK (53, 56, 57, 61), Brazil (17, 62), USA (64) and Canada (65). Differences within the group of Black HCWs must also be considered. For instance, data from the UK-REACH UK showed that vaccine hesitancy was lowest in the group of White British HCWs (21%) followed by White Other (29%), but it was much higher among Black Caribbean (54%) compared to Black African HCWs (33%) (57). An international report on nurses highlighted inequalities related to both race/ethnicity and migrant status (67). In Sweden, data illustrated the strongest disadvantage for HCWs from EU countries other than Sweden (66). German research looked more generally at migrant HCWs and revealed some disadvantages (45, 51, 52), but no coherent pattern could be identified.

Regarding the substance of inequalities, the scarcity of knowledge is even more serious. More than two third of the studies included in our analysis were concerned with just two major thematic areas: vaccination coverage and attitudes/hesitancy (55, 56, 57, 59, 60, 64), and infection and fear/risk of infection (51, 52, 53, 62, 63, 65). Disadvantages and discrimination were reported for all countries and both topics, except for one study in the UK that specifically investigated the persistence of vaccine hesitancy and did not identify significant differences related to an ethnic group (55). Other disadvantages for migrant HCWs were reported for personal protective equipment (PPE) (54, 65), mental health (58), and more generally higher levels of experienced inequalities (17, 61, 67).

#### What evidence is available on intersectionality and major categories of inequality?

Data collected in different countries highlighted that the disadvantages of migrant and racialised groups of HCWs during the pandemic strongly intersect with other inequalities, and may thus affect the migrant HCWF in different ways. Comparative research was able to identify a bundle of items that may intersect with the migration/racial status. The results showed that inequalities were created on all levels: the health system, the organisational settings, professional settings and individual conditions (Figure 1). Some selected examples illustrate the complexity of intersectional dynamics.

To begin with, ‘systemic racism’ was reported from a survey in the US (64). Qualitative studies carried out in the UK added further evidence that ‘institutional and structural racism’ created inequalities and discrimination in the HCWF (58, 59, 60). A multi-country nursing report brought the importance of new health labour market policy and welfare state arrangements into the debate, which were enhanced by the ‘redistribution of power relations’ during the pandemic (67).

Magri and colleagues (17), referring to Brazil, added in-depth information on intersecting dynamics enhanced by macro-level policy and healthcare governance. The authors argued that the conditions of the Brazilian National Health Service (SUS) ‘with its structural problems exacerbated by the recent precariousness and cut of recourses’ (17, p. 1439) determine what resources frontline HCWs have during the pandemic. They observed that some elements of discrimination ‘started from the professional characteristics of frontline workers’, yet professional careers are intertwined with ‘racial and gender markers’ that are therefore relevant to understanding the dynamics of individual reactions of HCWs (17, p. 1440).

Viewed through this intersectional lens, the research was able to explain, for instance, higher vulnerability to the conditions caused by the COVID-19 pandemic in the overlapping groups of Black women and community healthcare workers, who had less access to testing and training and felt less well-prepared than White men (17). A German study provided further evidence of the crucial intersections of health workforce governance, profession and gender, highlighting that the German system based on predominantly non-academic education of nurses has created inequalities for migrant nurses (45). Foreign-trained nurses often hold academic degrees and thus experience downgrading of their qualification in Germany.

In healthcare, professional and organisational settings are interconnected and inequalities created in one area may be reinforced in another. This was described, next to Brazil (17, 63) for Canada (65), Germany (45, 51, 52) and the UK (53, 54, 58, 61). For instance, disadvantages caused by lower PPE protection were strongest for lower status groups, e.g., hospital support worker (63), community healthcare workers (17), and HCWs working in residency care/long-term care (65). Yet disadvantages were also reported for those in dental compared to medical roles in the UK (54), thus reflecting health system specific hierarchies in the British NHS.

UK studies provided further examples of intersectionality. For instance, it was reported that migrant HCWs often work as agency nurses in the UK; agency workers are usually placed in higher-risk areas than other staff, thus facing higher risk of infection during the pandemic (61). To recall, the majority of nurses are women, and discrimination of agency work may reinforce gendered hierarchies in the HCWF. Another example was related to a stronger loss of trust in the employer during the pandemic in ethnic minorities, caused by due to the perception of institutional racism (58).

#### What is known on the German migrant HCWF?

Summing up the findings with a view on Germany, two studies focused on infection risk, reporting higher fear of being infected and infecting others among migrant HCWs (51) and higher risk for SARS-CoV-2 infection (52). A further study indicated that a migration background may be more precarious for nurses than for doctors due to the education system (Peppler and Schenk, 2022). All three studies highlighted intersectionality of inequalities, including age, sex, living conditions, working conditions, profession, and the education system.

### Comparing national-born and foreign-born HCWs in Germany

#### COVID-19-related health and psycho-social status

The sample drawn from the DEFEAT study comprised 1068 HCWs, of which 6.4% (n=68) had a migrant status. This figure is lower than the percentage of migrant HCWs in the HCWF, which might reflect language barriers, among others. However, the composition of the two groups of foreign-born and national-born HCWs was largely similar regarding gender, age and school education; no information was available on the profession. We did not find statistically significant differences concerning the health-related items included in the analysis: vaccination status, SARS-CoV-2 infection risk, pre-existing diseases, and health status. Yet a score related to social participation (IMET) pointed towards disadvantages of foreign-born HCWs (p 0.017) (Table 1).

**Table 1.** COVID-19 related health and psychosocial status, national-born and foreign-born HCWs

| Items | Total sample, n (%) | Foreign-born n (%) | National-born n (%) | P value |
| --- | --- | --- | --- | --- |
| Participants, n (%) | 1068 (100) | 68 (6) | 1000 (94) |  |
| Gender |  |  |  | 0.948<sup>1</sup> |
| Female | 910 (85) | 59 (87) | 851 (85) |  |
| Male | 154 (14) | 9 (13) | 145 (15) |  |
| Non-binary | 2 (0.2) | 0 | 2 (0.2) |  |
| Not answered | 2 (0.2) | 0 | 2 (0.2) |  |
| Age (years)/ median 25<sup>th</sup>–75<sup>th</sup> percentile | 42 (32–52) | 40 (32–47) | 42 (32–52) | 0.307<sup>2</sup> |
| Not answered | 1 | 0 | 1 |  |
| Education |  |  |  | 0.243<sup>1</sup> |
| High school | 711 (67) | 49 (72) | 662 (66) |  |
| Middle school | 328 (31) | 15 (22) | 313 (31) |  |
| Secondary school | 27 (3) | 4 (6) | 23 (2) |  |
| No school graduation | 1 (0.1) | 0 | 1 (0.1) |  |
| Not answered | 1 (0.1) | 0 | 1 (0.1) |  |
| Fully vaccinated* against SARS-CoV-2 |  |  |  | 0.365<sup>3</sup> |
| Yes | 833 (78) | 50 (74) | 783 (78) |  |
| No | 235 (22) | 18 (26) | 217 (22) |  |
| SARS-CoV-2 infection |  |  |  | 0.705<sup>3</sup> |
| Yes | 621 (58) | 38 (56) | 583 (58) |  |
| No | 447 (42) | 30 (44) | 417 (42) |  |
| Pre-existing diseases |  |  |  | 0.223<sup>3</sup> |
| Yes | 612 (57) | 36 (53) | 576 (58) |  |
| No | 417 (39) | 27 (40) | 390 (39) |  |
| Not answered | 39 (4) | 5 (7) | 34 (3) |  |
| EQ-5D Health status median<br>25<sup>th</sup>–75<sup>th</sup> percentile | 76<br>(52–91) | 76<br>(52–91) | 71<br>(51–92) | 0.908<sup>2</sup> |
| Not answered | 15 (1) | 3 (4) | 12 (1) |  |
| IMET median, 25<sup>th</sup>–75<sup>th</sup><br>percentile | 19<br>(8–41) | 31<br>(13–51) | 19<br>(7–40) | **0.017<sup>2</sup>** |
| Not answered | 57 (5) | 6 (9) | 51 (5) |  |

Source: DEFEAT study data (68), authors' own calculation
<sup>1</sup> Chi-square test; <sup>2</sup> Wilcoxon test; <sup>3</sup> Fisher Exact test
\* minimum two vaccinations

When unpacking the IMET score, the findings revealed strong disadvantages of migrant HCWs concerning the item ‘social activities’ (national born 3.514, SD 3.245 vs. foreign-born 4.768, SD 3.594; *p=0*.*004*), but not for ‘stress/burdens’ (national-born 4.403, SD 3.081 vs. foreign-born 5.029, SD 3.237; p=0.124).

#### Comparing perceptions of individual, organisational and government COVID-19 items

The sample drawn from the COVID-19 Contact study (CoCo) comprised a total of 468 participants. 428 (91.8%) were national-born; among the foreign-born HCWs (8.2%) 3.7% were EU foreign-born and 4.5% were non-EU foreign-born. Our comparative analysis did not reveal any significant differences in the composition of the groups concerning gender (75% women, 22% men, 3% not answered, total sample), age (43 years, mean), workplace (specified for emergency room, ward, intensive care unit/ICU, theatre room, ambulance, and others), and profession (specified for doctors, nurses, medical assistants, and others); there were also no relevant differences in relation to children and caring responsibilities. Notably, we also did not find statistically significant differences between EU foreign-born and non-EU foreign-born HCWs, except for a slightly lower mean age of non-EU foreign born participants.

Other relevant items selected for our analysis were grouped into five areas: health status, workplace conditions, perception of individual risks and behaviour, perception of organisational protection measures, and perception of the appropriateness of government pandemic measures. Major findings (Table 2) did not reveal a coherent pattern of inequalities for either of the groups of migrant HCWs. However, work place and task-related conditions put migrant HCWs at higher risk for COVID-19 infections compared to national-born HCWs, e.g., through more frequent contact with COVID-19 patients and more contacts without or with insufficient PPE. These items appeared to be statistically significant after merging the two groups of migrant HCWs. No significant differences could be identified between EU-foreign and non-EU foreign-born HCWs, except for exception of sickness leaves that were more often reported from non-EU born HCWs.

**Table 2.**
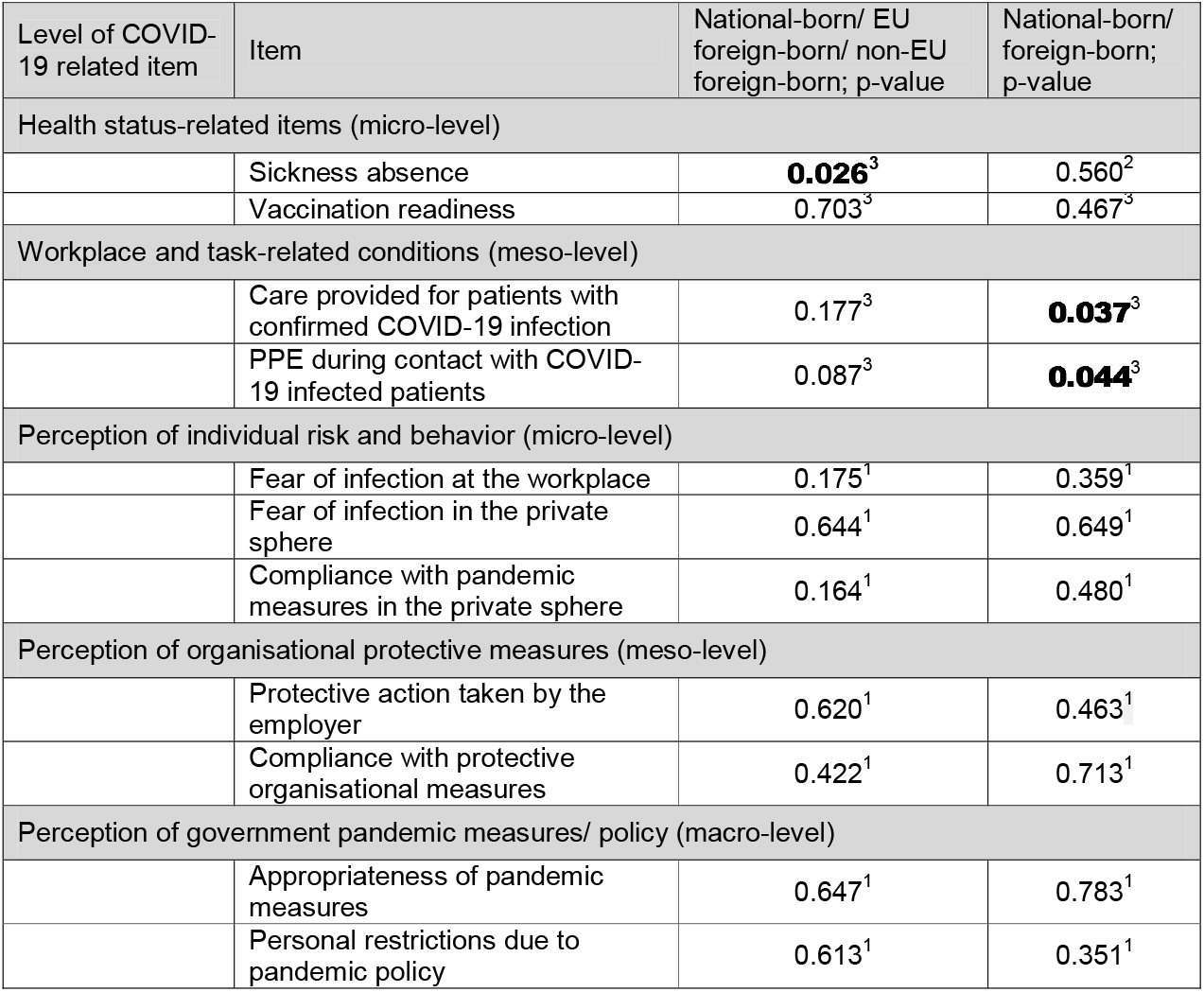
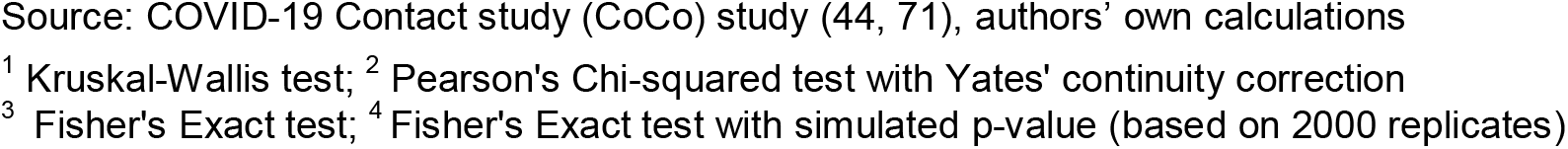
Perceptions of individual, organisational and governmental COVID-19 related items, comparison of national-born, EU foreign-born and non-EU foreign-born healthcare workers

| Level of COVID-19 related item | Item | National-born/ EU foreign-born/ non-EU foreign-born; p-value | National-born/ foreign-born; p-value |
| --- | --- | --- | --- |
| Health status-related items (micro-level) |  |  |  |
|  | Sickness absence | **0.026<sup>3</sup>** | 0.560<sup>2</sup> |
|  | Vaccination readiness | 0.703<sup>3</sup> | 0.467<sup>3</sup> |
| Workplace and task-related conditions (meso-level) |  |  |  |
|  | Care provided for patients with confirmed COVID-19 infection | 0.177<sup>3</sup> | **0.037<sup>3</sup>** |
|  | PPE during contact with COVID-19 infected patients | 0.087<sup>3</sup> | **0.044<sup>3</sup>** |
| Perception of individual risk and behavior (micro-level) |  |  |  |
|  | Fear of infection at the workplace | 0.175<sup>1</sup> | 0.359<sup>1</sup> |
|  | Fear of infection in the private sphere | 0.644<sup>1</sup> | 0.649<sup>1</sup> |
|  | Compliance with pandemic measures in the private sphere | 0.164<sup>1</sup> | 0.480<sup>1</sup> |
| Perception of organisational protective measures (meso-level) |  |  |  |
|  | Protective action taken by the employer | 0.620<sup>1</sup> | 0.463<sup>1</sup> |
|  | Compliance with protective organisational measures | 0.422<sup>1</sup> | 0.713<sup>1</sup> |
| Perception of government pandemic measures/ policy (macro-level) |  |  |  |
|  | Appropriateness of pandemic measures | 0.647<sup>1</sup> | 0.783<sup>1</sup> |
|  | Personal restrictions due to pandemic policy | 0.613<sup>1</sup> | 0.351<sup>1</sup> |

Source: COVID-19 Contact study (CoCo) study (44, 71), authors' own calculations
<sup>1</sup> Kruskal-Wallis test; <sup>2</sup> Pearson's Chi-squared test with Yates' continuity correction
<sup>3</sup> Fisher's Exact test; <sup>4</sup> Fisher's Exact test with simulated p-value (based on 2000 replicates)

In-depth information is provided for selected items. Figure 2 illustrates workplace and task-related conditions (based on the items ‘Care provided for patients with confirmed COVID-19 infection’ and ‘PPE during contact with COVID-19 infected patients’). Figure 3 shows how ‘fear of infection at the workplace’ was perceived, while Figure 4 draws on protective action taken by the employer, and Figure 5 on the appropriateness of government pandemic measures.

**Figure 2.**
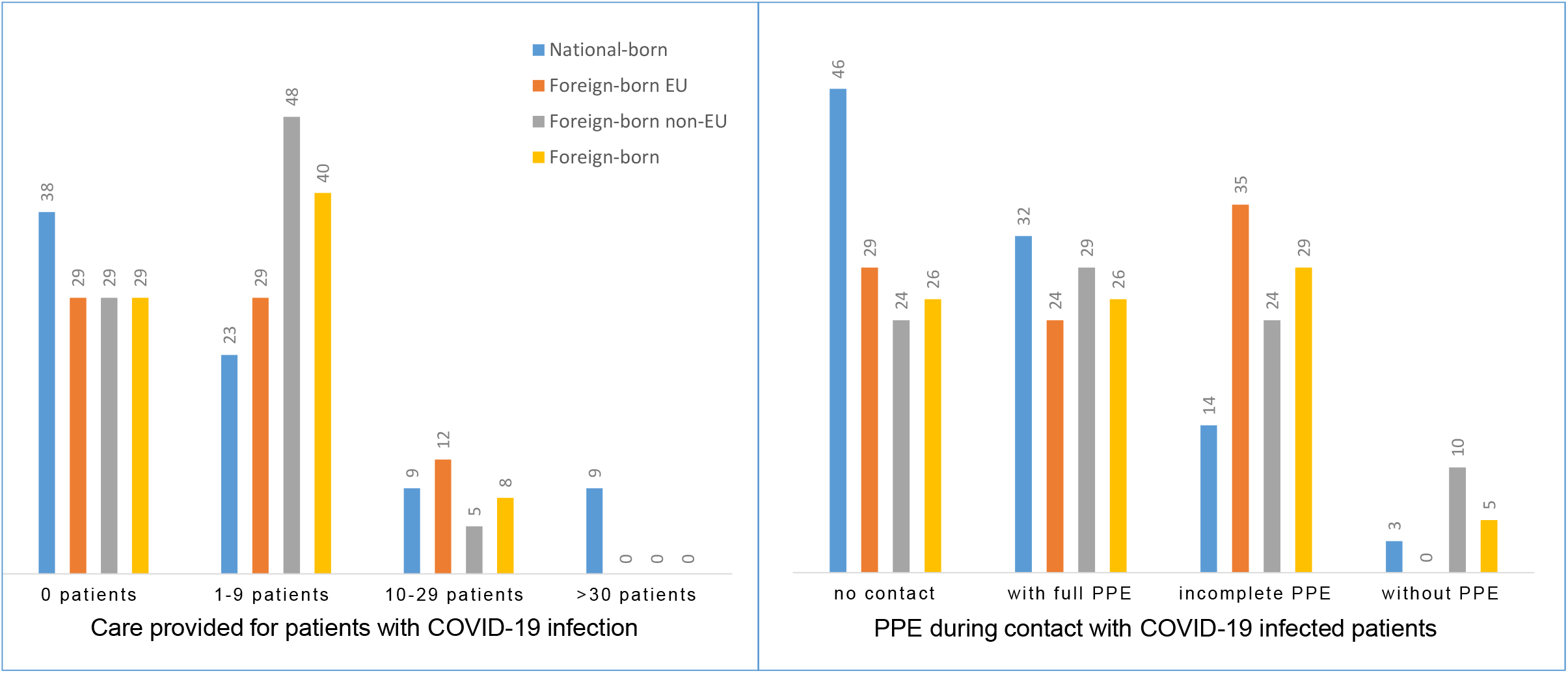
Workplace and task-related conditions (%) Source: Corona Contact study, authors’ own figure

**Figure 3.**
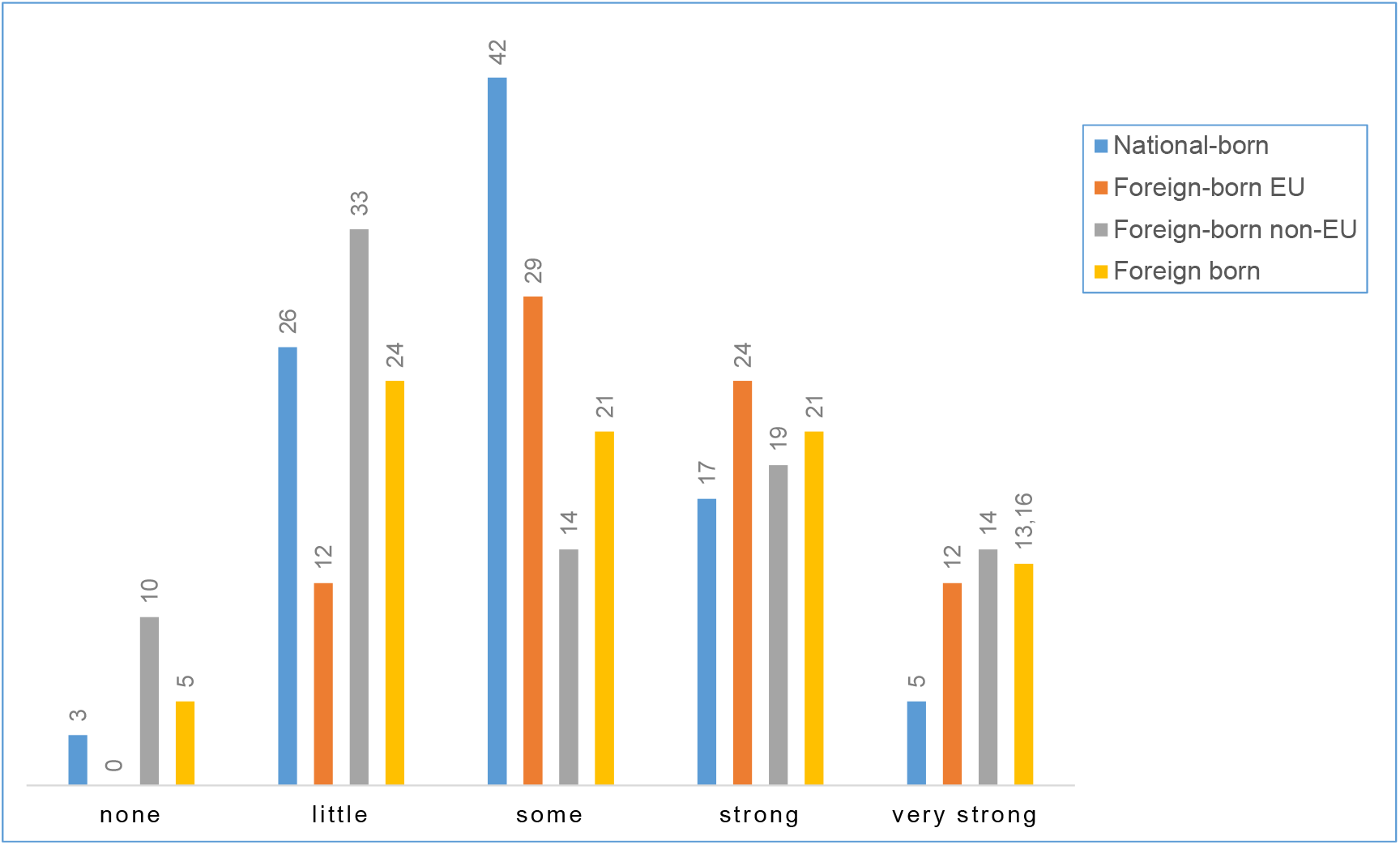
Fear of infection at the workplace, comparison (%) Source: Corona Contact study, authors’ own figure

**Figure 4.**
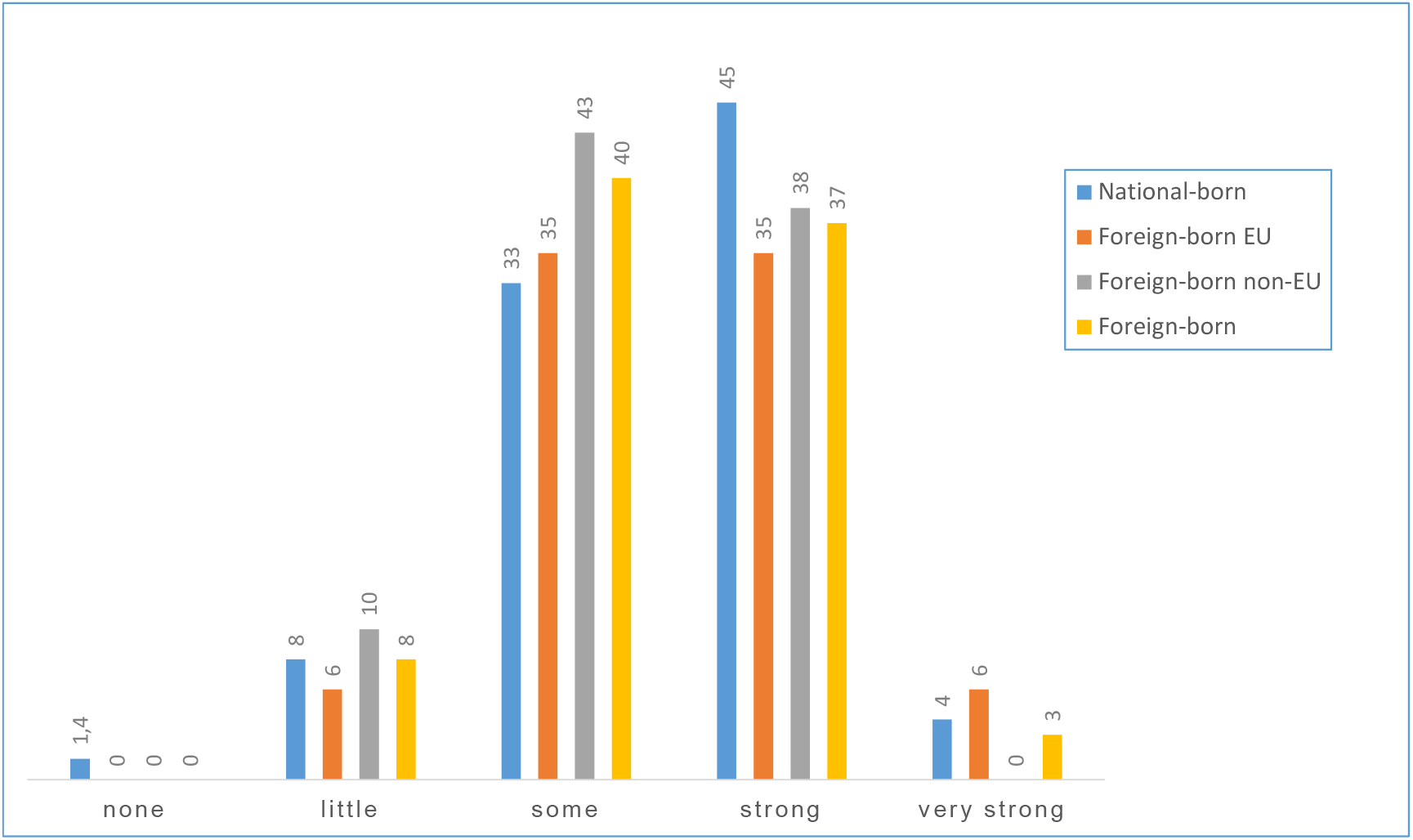
Protective action taken by the employer, perception (%) Source: Corona Contact study, authors’ own figure

**Figure 5.**
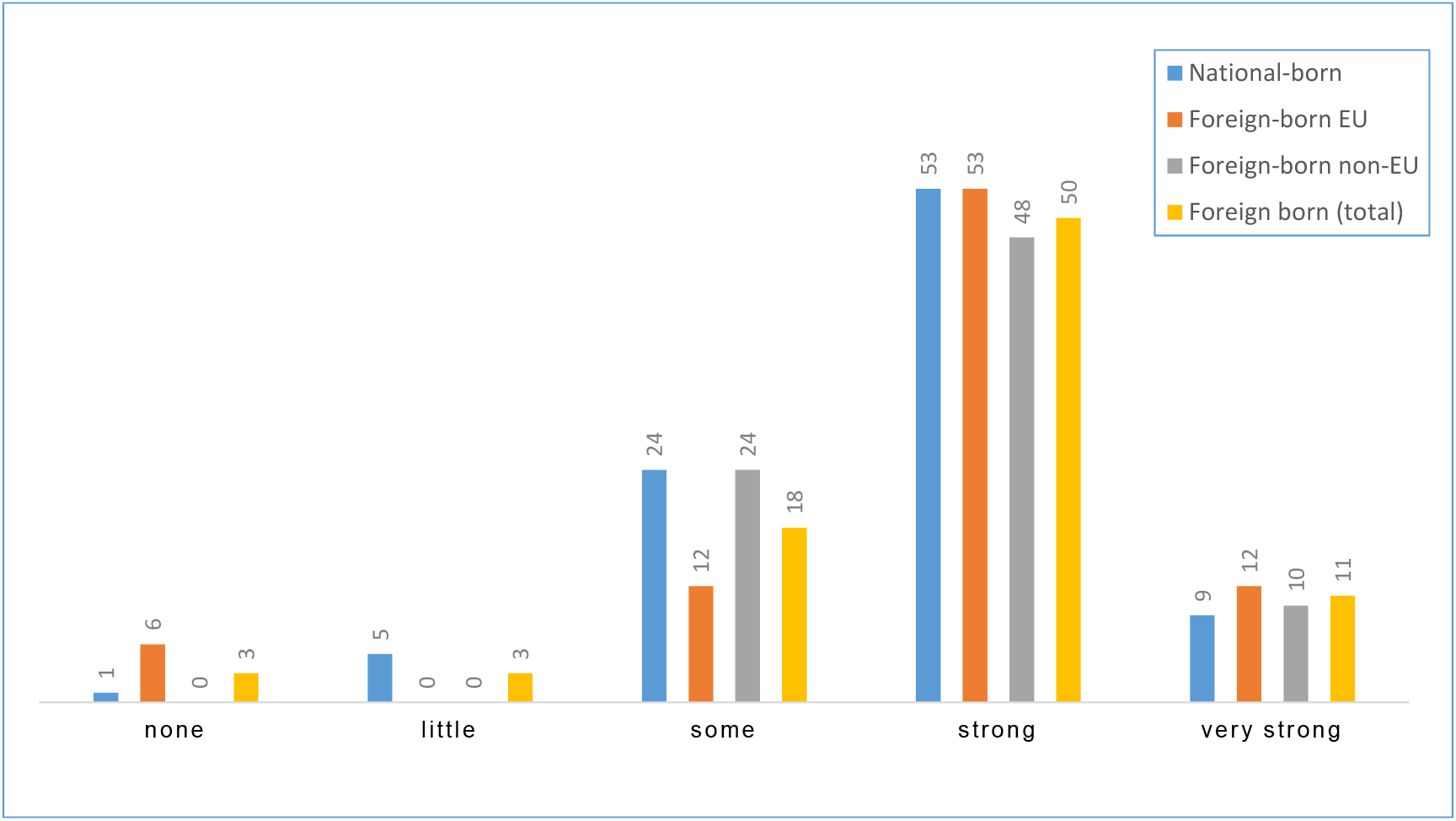
Agreement with government pandemic measures (%) Source: Corona Contact study, authors’ own figure

## Discussion

Our research highlighted an overall dearth of knowledge related to migrant HCWs during the COVID-19 pandemic, and data were especially poor for EU countries including Germany. This mirrors a general lack of attention to migrants and the social dimensions of the COVID-19 pandemic, despite early warnings of a clustering of risk factors according to ‘patterns of inequality deeply embedded in our societies’ (32, p. 874) and evidence of the gendered nature of these clusters, including its intersectional inequalities (13).

The findings confirmed inequalities and discrimination in the HCWF during the pandemic and we were able to specify some of the patterns of inequality. However, neither race/ethnicity nor the migration status measured through common categories of foreign-born or foreign-trained sufficiently explain inequalities. HCW migration and mobility are generally extremely difficult to measure and ‘cannot be described adequately with any single metric’, as a recent WHO report discussed in more detail (86, p. 204). Available evidence strongly calls for an intersectional approach. Our research framework, based on an intersectional health system-related approach, has proven useful to investigate the situation of migrant HCWs and consider different levels of health systems, organisations and professions, and individual actors. As such, we suggest that it can be further used and/or refined to explore in greater depth HCW migration and mobility.

The literature reported a range of inequalities created at different levels, that illustrate the complexity of the migrant status and the challenges of defining and analysing it. Intersecting inequalities do not simply sum up and can hardly be quantified (13, 17). However, the dynamics of inequalities seem to be particularly strong, if disadvantages created at different levels meet and the following major conditions apply:

- poor health system conditions and pandemic policy,
- lower status professional group,
- organisational settings with high infection risks and/or lack of effective pandemic protection including PPE,
- Black (and more generally, strong ‘othering’ and minority status, which may also put other groups at risk, depending on national contexts),
- female sex (information on non-binary/LGBTQ migrant HCWs was lacking).

Intersecting inequalities may explain why and how the COVID-19 pandemic exacerbated the situation of migrant HCWs in most countries more strongly than observed in the domestic HCWF, as our literature overview has illustrated. An intersectional analysis may also reveal positive dynamics. High-status professional groups and identity strongly grounded in professionalism emerged as major factors that mitigated (to a certain extent) the potentially negative effects of migration status. This benefits physicians the most and may thus reinforce existing hierarchies in the healthcare sector.

When looking at the German case, the disadvantages of migrant HCWs seemed to be much weaker compared to what was reported from the international literature. Indeed, in our empirical data sets, we rarely found any significant differences between national-born and foreign-born HCWs for most of the items. Notably, our data did not show significant differences for SARS-CoV-2 infection and vaccination status. For both items, the international literature highlighted strong disadvantages and structural and institutional racism. There are two important conclusions to be drawn from the results. Firstly, more favourable conditions may be explained by the fact, that most of the items previously identified as risk factors of exacerbated inequalities do not apply to our case study – it is important to recall here, that our research design was based on an optimum case scenario. To this end, the results highlight the importance of health systems, including meso-level conditions of organisations and professions.

The second important conclusion to be drawn from this case makes weaknesses and gaps in pandemic governance visible, especially on the organisational level. Migrant HCWs scored significantly lower for ‘social activities’ during the COVID-19 pandemic. This is important and may cause higher mental health risks, including the risk of burnout, as well as impede actions aimed at improving HCWs’ well-being (87). Since recently, attention to the mental health has improved (88, 89), including mental health of HCWs (90, 91), yet these efforts rarely consider the needs of migrant HCWs.

Furthermore, migrant HCWs were more often found in workplaces and tasks with higher numbers of COVID-19-infected patients and lower PPE protection. Similar conditions were described in the international literature. Bossavie and colleagues (37) asked whether ‘migrants shield the locals’ during the pandemic, and found that national workers in the EU move to lower-risk sectors/tasks and migrants take higher risk tasks (37). It is interesting to note, that we found statistical evidence for higher workplace risks for migrant HCWs in our research, but none of the items that measured individual perceptions, including protection by the employer, and health status did reflect differences, except higher sickness leaves in the group of non-EU foreign-born HCWs. A comparably low infection risk of HCWs at Hannover Medical School together with appropriate access to PPE and high vaccination coverage might explain this mismatch (31, 71).

### Limitations

Our study has several limitations, which are mainly related to the use of secondary data analysis, theoretical and methodological challenges of the research field, and scarcity of empirical data, in particular, lack of cross-country comparative research. Secondary data analysis naturally pre-defines the sample and available items, that may not perfectly match the research objectives.

One major limitation stems from the small subgroups of migrant HCWs in both the DEFEAT and CoCo study. These limitations were minimised, because for both studies we did not find relevant differences between the subgroups of HCWs created for our analysis for any of the items. However, small numbers naturally constrain statistical options; our analysis was therefore limited to descriptive statistics. Signs of disadvantages might be overlooked as small absolute numbers may not achieve statistical significance, while at the same time, a higher risk of statistical artefacts caused by a few out-layer cases must be considered. For instance, this might be the case for our finding of higher rates of sickness absence among non-EU foreign-born HCWs compared to EU foreign-born and national-born HCWs.

A further important limitation relates to common problems of migration studies: lack of data, lack of common definitions and categories, and appropriate methodological approaches (96). EU law of free movement of people (Greer et al., 2022) and cross-border mobility of HCWs add further challenges, turning research into migrant HCWs during an entirely novel situation of the COVID-19 pandemic into a walk on uncertain ground. We responded to these challenges in two ways. Firstly, we developed a generic methodological framework (Figure 1) that provided guidance and coherence throughout the research. Secondly, we applied a pragmatic approach, that considered different categories related to migration and race/ethnicity, and qualitative scoping review methodology (15, 48) to identify knowledge gaps and explore key themes. We may have overlooked information in the literature, that was not presented under one of the selected categories of migration and social inequalities.

The empirical data were mainly constrained through a focus on the category ‘foreign-born’, as other categories were not available. Importantly, the data drawn from DEFEAT and CoCo did not provide sufficient information or appropriate statistical options to analyse profession-specific effects. This is particularly relevant against the backdrop of strong intersections between the profession and inequalities, as identified in the literature. It is important to note that the research was designed as a pilot, aiming to highlight a need for better research evidence on migrant HCWs during the pandemic to inform policy and improve pandemic protection and health workforce governance.

## Conclusions and policy recommendations

This study set out to explore the situation of migrant HCWs during the COVID-19 pandemic, using Germany as a case study that reflects an optimum-case scenario in the European region. Three major conclusions and policy recommendations were emerging from our research.

- Investigation in comprehensive health labour market monitoring and comparative research into the situation of migrant HCWs is an urgent need to improve evidence and inform health policy and workforce governance.
- Simply adding the migration status is not enough. We introduced an intersectional health system-related research framework that revealed how health system and governance conditions played an important role in exacerbating the disadvantages of migrant HCWs, often along the lines of organisational settings and professional groups, including gendered hierarchies.
- The health labour market role and individual needs of migrant HCWs must be moved higher up on the policy agenda both nationally and globally. In a situation of a global health crisis, health system failures may impact like wildfire and spark destructive dynamics of existing inequalities stemming from the work and living conditions of migrant HCWs.

## Data Availability

ll data produced in the present study are available upon reasonable request to the authors

## Acknowledgments

We thank the project teams of the COVID-19 Contact study and the DEFEAT study at Hannover Medical School for making the data sets available for this secondary analysis.

## Ethics statement

The COVID-19 Contact (CoCo) study was approved by the Ethics Committee of Hannover Medical School, Nr. 8973_BO_K_2020. DEFEAT Corona was approved by the Ethics Committee of Hannover Medical School, Nr. 9948_BO_K_2021. Written informed consent was obtained from all subjects involved in the studies.

## Author contributions

EK and AD-J had the idea and supervised the research; EK, AJ-D and M-IU developed the design; EK prepared the draft, carried out the literature review and coordinated the analysis; AD-J supervised the statistical analyses; MM analysed the DEFEAT data; AC and SK analysed the CoCo data; M-IU and MGB advised on migrant healthcare workers; LMF, GMNB, FM, NT supported the data collection; all authors provided comments and have read and approved the final manuscript.

## Data availability statement

The raw data supporting the conclusions of this article will be made available by the authors, without undue reservation.

---

**Appendix Figure 1.**
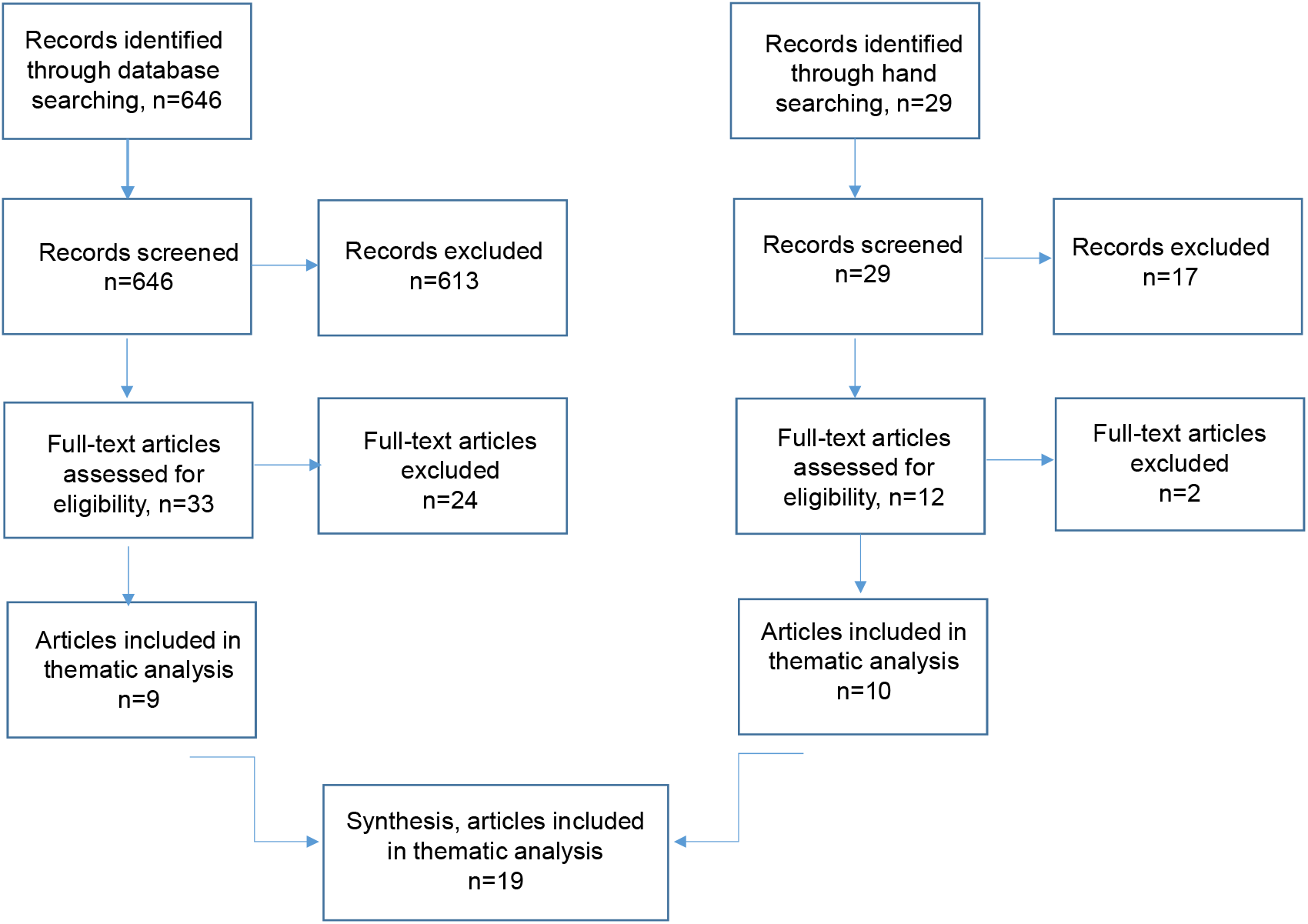
Flow chart scoping review

